# Global academic response to COVID-19: Cross-sectional study

**DOI:** 10.1101/2020.04.27.20081414

**Authors:** Jack A Helliwell, William S Bolton, Joshua R Burke, Jim P Tiernan, David G Jayne, Stephen J Chapman

## Abstract

**Objective:** To describe the global academic response to COVID-19 during its early stages. The responsiveness of investigators, editorial teams, and publishers was explored.

**Design:** Cross-sectional bibliometric review of COVID-19 literature. A parallel search of Middle East Respiratory Syndrome (MERS) literature was performed for comparison of outcomes.

**Data sources:** MEDLINE and EMBASE databases. The search for COVID-19 studies was performed between 1st November 2019 and 24th March 2020. The search for MERS studies was performed one year earlier between 1st November 2018 and 24th March 2019.

**Main outcome measures:** Investigator-responsiveness was assessed by measuring the volume and type of published research. Editorial-responsiveness was assessed by measuring the time from manuscript submission to acceptance and the availability of original data to support the study results. Publisher-responsiveness was assessed by measuring the time from manuscript acceptance to first publication and the provision of open access.

**Results:** In total, 398 of 2835 COVID-19 and 55 of 1513 MERS search results were eligible. Most COVID-19 studies were clinical reports (n=242; 60.8%) and the majority of these were case series (n=105; 43.4%) and single cases (n=65; 26.9%). The times from manuscript submission to acceptance (median: 5 days (IQR: 3–11) vs 71.5 days (38–106); *P*<0.001) and acceptance to publication (median: 5 days (IQR: 2–8) vs. 22.5 days (4–48.5-; *P*<0.001) were strikingly shorter for COVID-19. Almost all COVID-19 (n=396; 99.5%) and MERS (n=55; 100%) studies were available with open-access. Data sharing was infrequent, with original data available for 104 (26.1%) COVID-19 and 10 (18.2%) MERS studies (*P*=0.203).

**Conclusions:** The early academic response to COVID-19 was characterised by investigators aiming to define the disease. These studies were made rapidly and openly available by editorial and publishing teams. Data sharing practises are an essential target for improvement as the pandemic progresses.

## Introduction

Coronavirus disease 2019 (COVID-19) caused by the severe acute respiratory syndrome coronavirus 2 (SARS-CoV-2) has spread rapidly since it began in the city of Wuhan in late 2019[1]. The outbreak has been declared as a pandemic by the World Health Organisation (WHO), reflecting its spread to more than 200 countries worldwide[2]. As of April 2020, there has been more than 100,000 deaths and the number of those affected continues to rise[3].

In response to this unprecedented situation, healthcare systems around the world have taken urgent action to scale up medical staffing, equipment, and infrastructure. In the United Kingdom (UK), this is demonstrated by the construction of new field hospitals, the return of thousands of retired healthcare workers, and the re-purposing of many acute and urgent hospital services[4]. The role that science and data play in the face of the COVID-19 pandemic must also be recognised. Robust and rapidly-available evidence is essential for improving the detection and treatment of the virus as well as for reducing its transmission. It is the responsibility of all members of the academic community to facilitate this transfer of knowledge so that patients around the world receive the best evidence-based care available.

There is limited information about how the academic community has responded to COVID-19 and how this can be further guided during times of global crisis. There is a need for Investigators to respond urgently to new clinical needs across all sectors of healthcare and to work in unfamiliar environments. Journal editors must facilitate rapid and robust peer review whilst making difficult editorial decisions based on little previous academic background. Publishers must mobilise quickly to enable timely publication of manuscripts so that research outputs can be realised and implemented into practice. An early understanding of this response is important so that the academic community can identify early challenges and respond appropriately as the COVID-19 pandemic develops.

## Methods

### Ethics & Governance

As a review of published literature, approval by a research ethics committee was not applicable. Since only bibliometric outcomes were considered, rather than outcomes of direct relevance to research participants, this review was not eligible for registration on the PROSPERO database. There were no changes to the research design or outcomes during the course of the study. The results are reported with consideration to the Preferred Reporting Items for Systematic Reviews and Meta-Analyses (PRISMA) Checklist[5].

### Aims & Objectives

The study aimed to examine the responsiveness of the academic community to COVID-19 during its early stages. The following objectives were predefined:

1. To explore investigator-responsiveness by describing the volume and type of research accepted in peer-reviewed journals.
2. To explore editorial-responsiveness by describing the time taken to facilitate editorial/peer review and the availability of original data as a condition of publication
3. To explore publisher-responsiveness by describing the time taken for accepted manuscripts to be available and the provision of open access publication.

### Study Design

This was a systematic, cross-sectional, bibliometric review of existing literature related to COVID-19. To facilitate comparisons of outcomes in a unique setting, a comparable review of Middle East Respiratory Syndrome (MERS) was also performed and used as a non-COVID-19 control. MERS was chosen as a suitable comparator because it similarly represents a respiratory illness caused by a zoonotic coronavirus, but was not classified as a pandemic or a Public Health Emergency of International Concern by the WHO during the search period[6].

### Definitions

In line with the WHO, COVID-19 describes the disease caused by the severe acute respiratory syndrome coronavirus 2 (SARS-CoV-2) and MERS describes the disease caused by the Middle East respiratory syndrome-related coronavirus (MERS-CoV)[7]. The early stages of COVID-19 was defined as the period between the first known human case and its classification as a pandemic on 11^th^ March 2020[2]. Availability of original data was considered to be an editorial responsibility since this is recommended by the International Committee of Medical Journal Editors for clinical trials and can be enforced as a condition of publication[8]. Open-access was defined by the model of Gold open-access, where articles and related content are freely accessible at the point of publication. Fully open-access journals were defined by their listing on the Directory of Open Access Journals (ttps://doaj.org).

### Search Strategy

Systematic searches of MEDLINE (via OvidSP) and EMBASE (via OvidSP) were performed by a single investigator on 25^th^ March 2020. For COVID-19, time limits were set between 1^st^ November 2019 and 24^th^ March 2020. This start date was chosen to represent the earliest plausible time of the first human report. The end date reflected the official WHO classification of COVID-19 as a global pandemic (11^th^ March 2020) along with a 14-day lag period for manuscripts already in production. For MERS, a comparable search was performed between 1^st^ November 2018 and 24^th^ March 2019. The difference in year was chosen to represent a non-COVID-19 control period. The results of both searches were first deduplicated and saved offline for inspection. Two independent investigators then screened titles, abstracts, and full-texts for possible inclusion, with discrepancies addressed through re-examination and discussion. A full outline of search strategies is available in Suppl. Table 1.

### Study Eligibility Criteria

For published studies, any COVID-19- or MERS-related manuscript available online or in print within the defined time periods was eligible for inclusion. Eligible studies had to report primary data. This included studies which performed analyses using open source datasets, but not studies which reported systematic reviews or meta-analyses. Studies of animals or pre-clinical models were eligible, but only if the outcomes contributed to human rather than veterinary medicine. Articles published in non-English languages and other grey literature (such as conference abstracts) were excluded since these are not practically accessible by the global academic community.

### Study Outcomes

Academic responsiveness was considered across three groups of the academic community. Investigator-responsiveness was explored by measuring the volume and type of research (pre-clinical, clinical, modelling) published in peer-reviewed journals. Editorial-responsiveness was explored by measuring the time taken for editorial/peer-review, defined as the number of days between manuscript submission and acceptance. The availability of original data to support the results was also explored. Data were considered available if individual observations (or source code in the case of modelling studies) were provided as a supplementary file, in a controlled-access repository, or if an explicit statement of availability from authors was present. Where the status of data sharing was unclear, corresponding authors were contacted and non-responses were considered to represent an absence of data. Publisher-responsiveness was explored by measuring the time taken for manuscripts to complete production, defined as the number of days between acceptance and first availability online or in print. Publisher responsiveness was also assessed by exploring the incidence of open access publication.

### Data Collection

Data were collected by a single investigator and checked by an independent investigator for accuracy. Data sources included full-text manuscripts and editorial-related content. Data points of interest were country of origin (according to the corresponding institution), manuscript format (short communication/letter or full-text), journal subject category, and subject category ranking (according to Thomas Reuters Journal Citation Reports). For clinical studies, additional data of interest were population (adults, children, pregnant adults, or healthcare workers), study design (case report, case series, observational, or interventional) and clinical focus (definition of disease, diagnosis/screening, treatment, prevention, or resource use). Definition of disease related to studies describing clinical features and outcomes.

### Statistical Analysis

Data are presented descriptively using averages and measures of variance. Continuous data were analysed using the student’s t-test or analysis of variance (ANOVA). Categorical data were analysed using χ^2^ or Fisher’s Exact test. All statistical comparisons of COVID-19 and MERS studies were pre-planned. A single sub-analysis of data sharing practices was performed with the exclusion of case reports since these may or may not provide the full complement of data within the reported manuscript. The level of statistical significance was set at *P*<0.05.

## Results

### Study Characteristics

Across both searches, 398 of 2835 COVID-19 search results and 55 of 1513 MERS search results were eligible for inclusion (Figure 1 & Figure 2). The majority of COVID-19 studies were authored from institutions in China (n=254; 63.8%) whereas the majority of MERS studies were authored from the United States (n=14; 25.5%). A greater proportion of COVID-19 studies were published as letters or short communications (n=158; 39.7%) compared to MERS (n=4; 7.3%). Infectious Diseases (n=117; 29.4%) and General & Internal Medicine (n=78; 19.6%) were the most common journal subject categories for COVID-19, with the majority (n=248; 62.3%) of manuscripts published in the first quartile of category-specific rankings. In contrast, the most common category for MERS studies was Virology (n=15; 27.3%) (Table 1).

**Figure 1.**
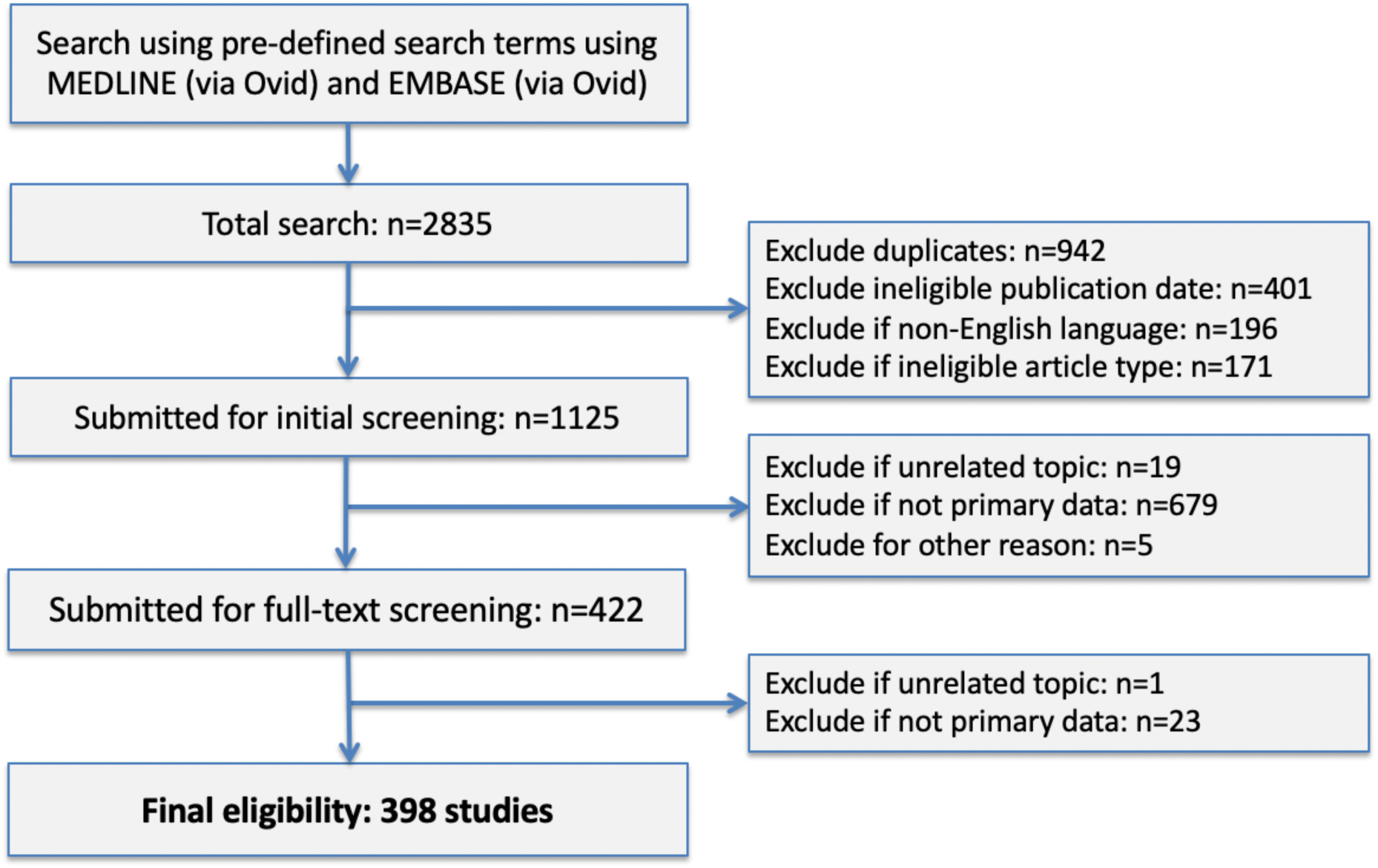
Flow diagram of COVID-19 eligibility

**Figure 2.**
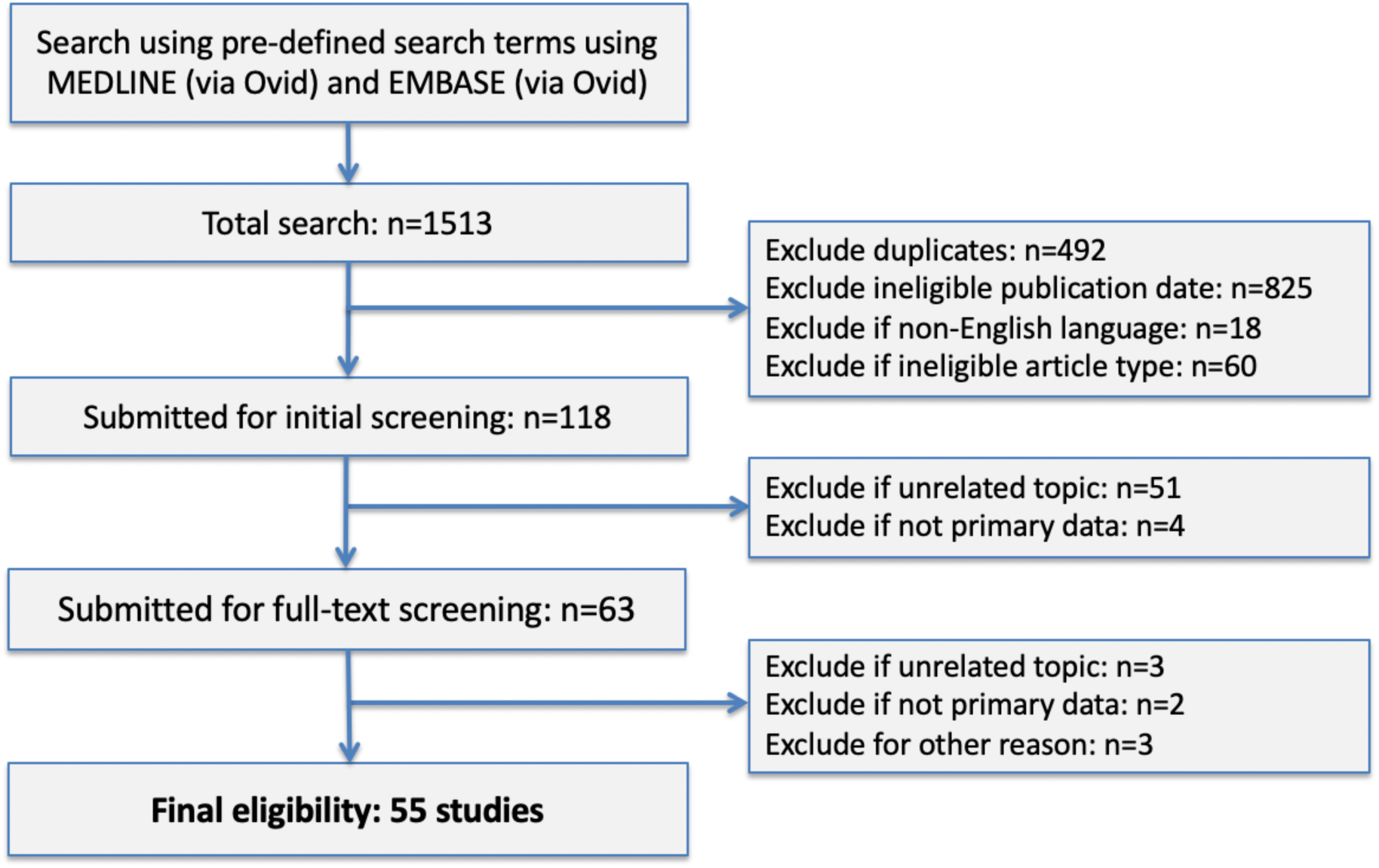
Flow diagram of MERS eligibility

**Table 1.** Bibliometric characteristics of COVID-19 and MERS published studies

|  |  | COVID-19<br>(n=398) | MERS<br>(n=55) |
| --- | --- | --- | --- |
| Country of publication* | China | 254 (63.8%) | 7 (12.7%) |
|  | USA | 28 (7.0%) | 14 (25.5%) |
|  | Japan | 13 (3.3%) | 0 (0.0%) |
|  | Korea | 12 (3.0%) | 12 (21.8%) |
|  | Singapore | 10 (2.5%) | 0 (0.0%) |
|  | Canada | 9 (2.3%) | 0 (0.0%) |
|  | Italy | 9 (2.3%) | 0 (0.0%) |
|  | Germany | 8 (2.0%) | 4 (7.3%) |
|  | UK | 8 (2.0%) | 0 (0.0%) |
|  | Saudi Arabia | 0 (0.0%) | 8 (14.5%) |
|  | Others† | 47 (11.8%) | 10 (18.2%) |
| Format of publication | Letter/ communication | 158 (39.7%) | 4 (7.3%) |
|  | Full-text manuscript | 240 (60.3%) | 51 (92.7%) |
| Type of Research | Pre-clinical | 90 (22.6%) | 38 (69.1%) |
|  | Clinical | 242 (60.8%) | 13 (23.6%) |
|  | Modeling | 62 (15.6%) | 1 (1.8%) |
|  | Other/miscellaneous‡ | 4 (1.0%) | 3 (5.5%) |
| Journal Category§ | Infectious Diseases | 117 (29.4%) | 10 (18.2%) |
|  | Medicine, General & Internal | 78 (19.6%) | 1 (1.8%) |
|  | Radiology, Nuclear Medicine & Medical Imaging | 44 (11.1%) | 0 (0.0%) |
|  | Virology | 39 (9.8%) | 15 (27.3%) |
|  | Microbiology | 16 (4.0%) | 6 (10.9%) |
|  | Multidisciplinary Sciences | 8 (2.0%) | 3 (5.5%) |
|  | Others** | 96 (24.1%) | 20 (36.4%) |
| Journal Category Quartile | First | 248 (62.3%) | 20 (36.4%) |
|  | Second | 50 (12.6%) | 19 (34.5%) |
|  | Third | 79 (19.8%) | 4 (7.3%) |
|  | Fourth | 5 (1.3%) | 4 (7.3%) |
|  | Unclassified | 16 (4.0%) | 8 (14.5%) |

\*Determined according to the corresponding institution
† All others n<6 including: Australia, Belgium, Egypt, France, Greece, Hong Kong, Hungary, India, Mexico, Nepal, Netherlands, New Zealand, Pakistan, Spain, Sweden, Switzerland, Taiwan, Thailand, Vietnam
‡ All miscellaneous studies were surveys; § According to Thomas Reuters Journal Citation Reports; †† Ranking according to category-specific Impact Factor
\*\* All others n<8 including: Anesthesiology; Biochemical Research Methods; Biochemistry & Molecular Biology; Biology; Cell Biology; Chemistry, Medicinal; Chemistry, Analytical; Critical Care Medicine; Dentistry, Oral Surgery & Medicine; Dermatology; Electrochemistry; Environmental Sciences; Gastroenterology & Hepatology; Genetics & Heredity; Hematology; Immunology; Medical Laboratory Technology; Medicine, Research & Experimental; Neurosciences; Oncology; Pediatrics; Pharmacology & Pharmacy; Psychiatry; Public, Environmental & Occupational Health; Respiratory System; Surgery; Unclassified

### Investigator Responsiveness

The first eligible COVID-19 study was reported on 21st January 2020. The volume of reports increased rapidly during the early stages of the disease, with 23 published in January 2020 (5.8%), 169 in February 2020 (n=42.5%), and 206 in March 2020 (51.8%). This compared to a median of 9 MERS studies per month (range: 5–20) published during the control period (Figure 3). The majority of COVID-19 studies were clinical (n=242; 60.8%), followed by pre-clinical (22.6%), and modeling studies (15.6%). Case reports (n=65; 26.9%) and case series (n=105; 43.4%) were the predominant designs for clinical studies and most of these set out to define the disease (n=125; 51.7%). Almost all clinical studies explored general adult populations (n=209; 86.4%), with a handful exploring other groups, including children (n=16; 6.6%), pregnant adults (n=7; 2.9%) and healthcare workers (n=10; 4.1%) (Table 2). In contrast, MERS studies were mostly pre-clinical (n=37; 67.3%), followed by clinical (n=13; 23.6%), modeling (n=1; 1.8%) and other miscellaneous (n=3; 5.5%). MERS clinical studies mainly comprised of case series and observational designs (both n=5; 38.5%) and most set out to define the disease or explore issues of diagnosis/screening (both n=5; 38.5%). The majority explored general adult populations (n=11, 84.6%).

**Figure 3.**
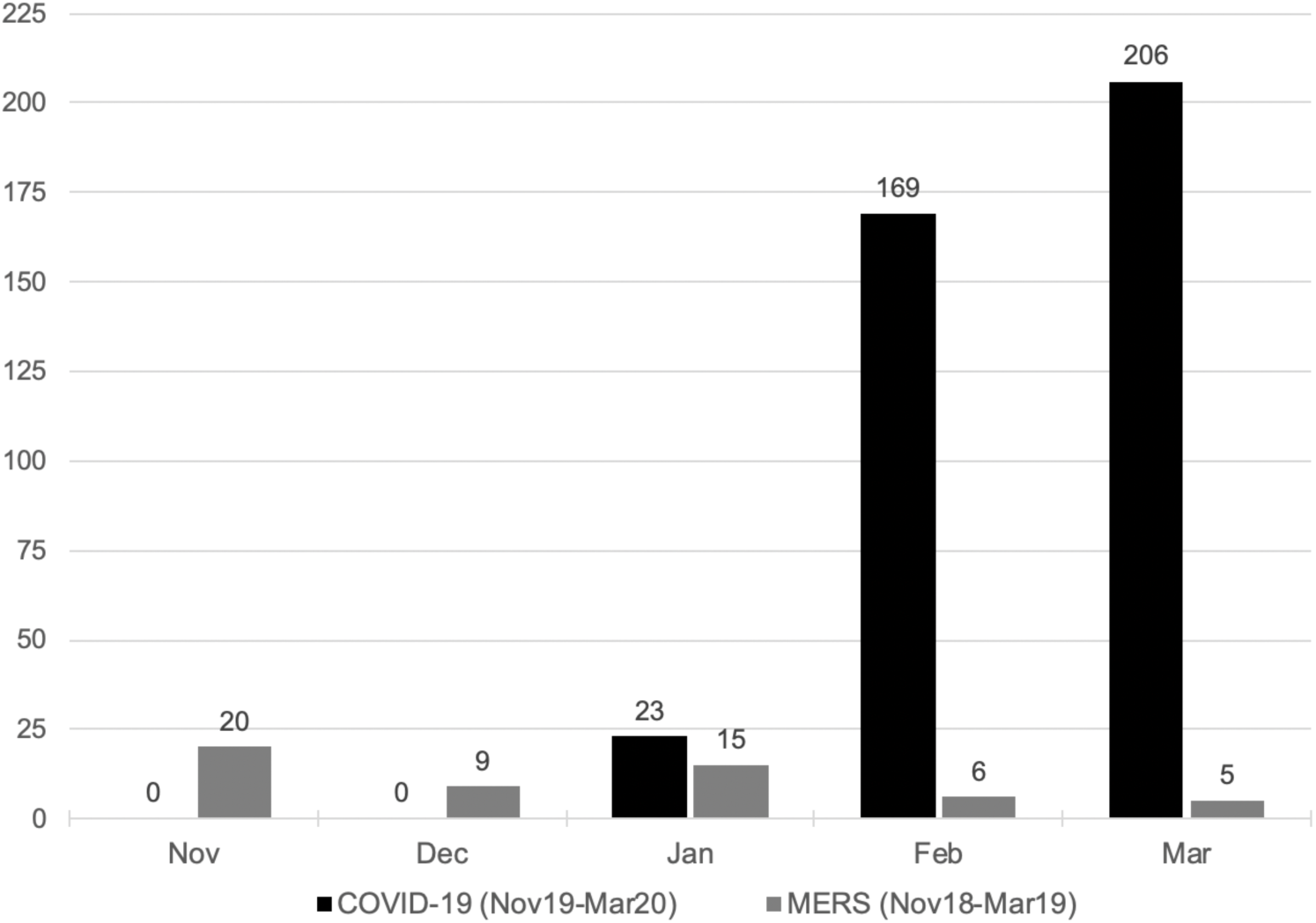
Volume of COVID-19 and MERS studies by month of publication

**Table 2.** Characteristics of clinical COVID-19 and MERS studies

|  |  | COVID-19 Published<br>(n=242) | MERS Published<br>(n=13) |
| --- | --- | --- | --- |
| Study population | Adults | 209 (86.4%) | 11 (84.6%) |
|  | Children | 16 (6.6%) | 0 (0.0%) |
|  | Pregnant adults | 7 (2.9%) | 0 (0.0%) |
|  | Healthcare workers | 10 (4.1%) | 2 (15.4%) |
| Study design | Case Report | 65 (26.9%) | 1 (7.7%) |
|  | Case Series | 105 (43.4%) | 5 (38.5%) |
|  | Observational | 64 (26.4%) | 5 (38.5%) |
|  | Interventional | 5 (2.1%) | 2 (15.4%) |
|  |  | Novel conjunctival secretion RT-PCR test | - |
|  |  | Novel rapid IgM-IgG antibody test | - |
|  |  | Ribavirin, interferon-alpha, lopinavir/ritonavir | - |
|  |  | Oxygenation-assisted tracheal intubation | - |
|  |  | Lopinavir/ritonavir | - |
|  | Other | 3 (1.2%) | 0 (0.0%) |
| Study focus | Definition of disease | 125 (51.7%) | 5 (38.5%) |
|  | Diagnosis/screening | 81 (33.5%) | 5 (38.5%) |
|  | Prevention | 15 (6.2%) | 0 (0.0%) |
|  | Treatment | 13 (5.4%) | 3 (23.1%) |
|  | Resource use | 1 (0.4%) | 0 (0.0%) |
|  | Other miscellaneous | 6 (2.5%) | 0 (0.0%) |

*RT-PCR: real-time reverse transcription polymerase chain reaction*

### Editorial Responsiveness

The time taken for editorial- and peer-review was available for 257 (64.6%) COVID-19 studies and 52 (94.5%) MERS studies. The median time from submission to acceptance was much shorter for COVID-19 studies (median: 5 days; interquartile range (IQR): 3–11) compared to MERS (median: 71.5 days; IQR: 38–106) (*P*<0.001). The median time for COVID-19 short communications was shorter than for full-texts (4 days, IQR: 2–7.5 vs. 6 days, IQR 3–13; *P*<0.001), whereas no significant difference in time was found between study designs (case report: 4.5, IQR 2–10 vs. case series: 5, IQR 2.25–10 vs. observational: 6, IQR 3–13; *P*=0.645). Original data were available via a supplement, data repository, or through direct request from the study authors for 104 (26.1%) COVID-19 studies compared to 10 (18.2%) MERS (*P*=0.203). When case reports were excluded, data were available for 95 of 333 (28.5%) COVID-19 studies and 10 out of 54 (18.5%) MERS studies.

### Publisher Responsiveness

The time taken for production processes was available for 280 (70.4%) COVID-19 and 52 (94.5%) MERS studies. The median time from acceptance to first publication was significantly shorter for COVID-19 (5 days, IQR: 2–8) compared to MERS (22.5 days, IQR: 4–48.5; *P*=0.001). There was no significant difference in the time for production between letters/short communications (median: 5, IQR: 1.5–8) and full-texts (median: 5, IQR: 2–8) (P=0.617). Neither was there a significant difference in production time between study designs (case report: 5, IQR: 3–8 vs. case series: 5, IQR 2–8 vs. observational: 5, IQR: 3–7.75) (*P*=0.954). A total of 396 (99.5%) COVID-19 manuscripts were available with open-access, including 275 (69.4%) manuscripts published in hybrid journals (open-access not mandated). Unexpectedly, all 55 (100%) MERS manuscripts were also available with open access, including 26 (47.3%) manuscripts published in hybrid journals. The difference in open-access between the two diseases was not significant (*P*=1.000).

## Discussion

The results demonstrate a rapid academic response to COVID-19 during its early stages. Investigators initially responded with case reports and case series in an attempt to describe the disease. These were highly published, often in major general medical or highly ranked specialty journals. The volume of interventional studies exploring treatments was low, but this is expected to change as ongoing trials reach completion. The editorial and production times for COVID-19 studies were strikingly shorter compared to MERS controls and almost all manuscripts were openly accessible at the point of publication. In contrast, only one-in-four COVID-19 manuscripts were published with original data available to support the results.

During times of medical crisis and global disease, rapid dissemination of information is essential to inform frontline treatments and prevention. This review provides an early insight into the challenges of the COVID-19 academic response. Even from the early stages, it is evident that data sharing practices are lacking and are a target for improvement as the pandemic progresses. A key strength of this review is the ability to identify this promptly and inform the study design, data management, and consent procedures of future research. A weakness of the review is the incomplete representation of the research pathway. Whilst investigators, editors, and publishers are essential for generating and disseminating research, other groups such as funding bodies, research ethics committees, regulatory agencies, and patient/stakeholder groups also play important roles. Measuring the response of these groups, however, is challenging since it relies on data which are not openly available or straight forward to measure. Another weakness is the focus on peer-reviewed publications. It is likely that a wider body of unpublished data exist following editorial rejection or through investigators making results available on non-peer reviewed platforms. These are important data, but it is proposed that further work to confirm their rigor and stability (i.e. changes made during parallel submission and peer-review) is required before wider consideration.

The academic response during times of global disease has been explored in the past. In a bibliometric analysis of the 2003 severe acute respiratory syndrome (SARS) crisis, the difference in median submission-to-acceptance intervals between SARS and non-SARS articles was 106.5 days (95% CI 55.0 to 140.1) and the difference in median acceptance-to-publication intervals was 63.5 days (18.0 – 94.1) [9]. In another bibliometric review of the West African Ebola Virus Disease (EVD), the academic response was shown to be organised around a small number of individuals with extensive global networks. This demonstrated the role and importance of strategic planning through international cooperation and expertise[10]. More recently, two early analyses of COVID-19 research found that most data were generated from China and called for increased academic output, particularly in the form of interventional trials of new treatments[11,12]. As suggested by the previous EVD response and the current data, this must be done with global coordination and in the spirit of open science. As more studies emerge during the COVID-19 pandemic, data sharing will be critical for allowing scrutiny of results, reducing unnecessary duplication, and enabling rigorous pooled analyses.

This study raises important considerations for the dissemination of data during the COVID-19 pandemic. Firstly, the urgency for information and the need for rigorous peer review must be balanced, particularly in studies that are controversial, preliminary, or practice-changing. Preprint publication is an alternative to fast-track editorial processes provided that audiences can accept the lack of prior editorial review[13]. Secondly, open-access publication is essential so that the global medical community can learn freely from each other. In this study, almost all articles were available with open-access for both COVID-19 and MERS research. It is likely that this represents and extended effort by publishers to share knowledge, since only 45% of the scholarly literature in 2015 was published with open-access[14]. Finally, in an age of electronic dissemination, it is clear that journals are not the only platform for reporting research data. International bodies such as the WHO publish regular COVID-19 situation reports and social media has emerged as a rapid source of knowledge transfer[3,15]. There remains a need to balance pragmatic and timely dissemination of results (not just limited to academic journals) with the assurances and validation brought by traditional peer-review.

There is no question that the academic response to COVID-19 from investigators, editors, and publishers has been prompt. Data sharing is an early unmet challenge and a commitment from all members of the academic community is required. Secure access data repositories exist to facilitate this safely, as do guidelines for data stewardship and management[16]. As the pandemic develops, it is likely that the anatomy of COVID-19 research will shift from efforts to describe the disease towards efforts to treat and prevent it. As with previous global diseases, a research agenda will be essential to ensure the global response remains coordinated and relevant[17]. Further auditing of the academic response will be important so that the academic community can identify and address new challenges as they emerge.

## Data Availability

All available data will be available after peer-reviewed publication

---

**Suppl. Table 1.** COVID 19 and MERS search strategies (via OvidSP) (24th March 2020)

|  |  |
| --- | --- |
| COVID-19 |  |
| 1 | SARS-CoV-2 |
| 2 | nCoV-19 |
| 3 | 2019-nCoV |
| 4 | COVID-19 |
| 5 | Novel coronavirus |
| 6 | Severe acute respiratory syndrome coronavirus 2 |
| 7 | 1 OR 2 OR 3 OR 4 OR 5 OR 6 |
| 8 | [Remove duplicates] |
| 9 | [Time limit: 2019-Current] |
| 10 | [Limit to English] |
| MERS |  |
| 1 | MERS |
| 2 | MERS-CoV |
| 3 | Middle East Respiratory Syndrome |
| 4 | 1 OR 2 OR 3 |
| 5 | [Time limit: 2018-2019] |
| 6 | [Limit to English] |
| 7 | [Remove duplicates] |

